# Programmatic diagnostic accuracy and clinical utility of Xpert MTB/XDR in patients with rifampicin-resistant tuberculosis in Georgia

**DOI:** 10.1101/2024.10.02.24314770

**Authors:** Theresa Pfurtscheller, Ana Tsutsunava, Nino Maghradze, Mariam Gujabidze, Nino Bablishvili, Seda Yerlikaya, Claudia M. Denkinger, Nestani Tukvadze, Ankur Gupta-Wright

**Author notes:** Corresponding Author: Theresa Pfurtscheller, Department of Infectious Diseases and Tropical Medicine, University Hospital Heidelberg, Heidelberg, Germany. contributed equally (Nestani Tukvadze, Ankur Gupta-Wright).

## Abstract

**Background:** Xpert MTB/XDR (Cepheid, USA) is recommended for drug susceptiblity testing in patients with tuberculosis (TB) by the World Health Organization (WHO) with potential for rapid detection of isoniazid and fluoroquinolones resistance. However, diagnostic accuracy and clinical utility in a programmatic setting are unknown.

**Methods:** We evaluated accuracy and clinical utility of Xpert XDR in patients with rifampicin-resistant pulmonary TB during programmatic implementation in Georgia between July 2022 and August 2024, using phenotypic drug susceptibility testing (pDST) as reference standard.

**Results:** 140 patients were tested with Xpert MTB/XDR and pDST, and 94.9% and 33.8% had isoniazid and fluoroquinolone resistance by pDST respectively. Xpert MTB/XDR showed 99.2% sensitivity (95% CI 95.5-100%) and 100% specificity (95% CI 54.1-100%) for isoniazid resistance. Sensitivity and specificity for fluoroquinolone resistance were 88.4% (95% CI 74.9-96.1%) and 100% (95% CI 95.6-100%). Including indeterminate/invalid Xpert MTB/XDR results, 17.4% (8/46) and 6.4% (8/129) of patients with phenotypic fluoroquinolone and isoniazid resistance were missed, respectively. Median turn-around time for Xpert MTB/XDR was 1 day (IQR 1-3) and median time to treatment was 4 days (IQR 1-7). Phenotypic DST results took a median of 43 days (IQR 29-63) longer than Xpert MTB/XDR results. 95% (115/121, 95% CI 89.5-98.2%) of patients had fluoroquinolones appropriately prescribed based on Xpert XDR result.

**Conclusions:** Progammatic data confirms high accuracy of Xpert MTB/XDR, although below WHO TPP targets for fluoroquinolones, with significantly faster time-to-results than pDST. However, there is a strong need for rapid tests to detect resistance to bedaquiline and other newer TB drugs.

## Background

Drug-resistant tuberculosis (DR-TB) remains a global health concern with an estimated 410,000 cases and up to 85,000 deaths attributed annually [1, 2]. Diagnosis remains a key barrier, with only 43% of those with drug-resistance starting appropriate treatment, of whom only 63% achieve favourable treatment outcomes [1]. Furthermore, selecting effective treatment requires identifying resistance to key drugs including rifampicin, isoniazid, and fluoroquinolones. The mainstay of drug susceptibility testing (DST) for TB is culture-based, with long turn-around-times, the need for expensive laboratory infrastructure, and associated costs, making implementation challenging [3]. The development and scale-up of rapid tests for resistance detection in correspondence to new treatment regimens is a priority for the World Health Organization (WHO) [4].

In 2021, the WHO recommended Xpert MTB/XDR (Xpert XDR, Cepheid, USA), a rapid, low-complexity molecular assay, as a follow-on test for detection of isoniazid and fluoroquinolone resistance in patients with bacteriologically confirmed pulmonary TB [5]. Xpert XDR demonstrated high diagnostic accuracy for isoniazid (sensitivity 94.2%, specificity 98.5%) and fluoroquinolone (sensitivity 93.2%, specificity 98.0%) resistance in multi-centre diagnostic accuracy studies [6-9].

Different programmatic use-cases have been suggested for Xpert XDR. In presumptive TB patients with high risk for resistance, it was suggested as a test for TB diagnosis and resistance detection. Additionally, in patients with rifampicin-resistant TB and patients not responding to treatment, it could facilitate detection of resistance to first- and second-line drugs [7]. While the relevance of second-line injectable drug resistance on Xpert XDR is less following the scale-up of all-oral treatment regimens for DR-TB, Xpert XDR remains valuable as the only rapid test available for the detection of resistance to isoniazid and fluoroquinolones at lower levels of care. However, its diagnostic accuracy in a programmatic setting, as well as its utility in clinical management of DR-TB patients, is not well established. We sought to evaluate the accuracy and clinical utility of Xpert XDR in a setting with high prevalence of DR-TB when implemented within routine programmatic care.

## Methods

### Study Design, Setting, and Participants

We evaluated diagnostic accuracy of Xpert XDR implemented programmatically at the National Centre for TB and Lung Disease (NCTLD) in Tbilisi, Georgia between July 4, 2022 and August 1, 2024. Results from programmatic testing of sputum, including Xpert MTB/RIF Ultra, Xpert XDR, mycobacterial culture, and phenotypic DST (pDST) were prospectively collected from consecutive patients with rifampicin-resistant TB enrolled in the Rapid Research in Diagnostics Development (R2D2) TB Network study on rapid DST assays for DR-TB [10]. Eligibility criteria for the R2D2 DR-TB study included rifampicin-resistant pulmonary TB, aged ≥12 years (≥18 years until 8^th^ September 2023), providing study sputum samples, and not being on effective DR-TB treatment for >72 hours. Patients without growth of *Mycobacterium tuberculosis* (MTB) complex on at least one sputum culture were excluded. Patients without a valid Xpert XDR result were excluded from programmatic diagnostic accuracy analysis, although they were included for clinica utility analysis. Eligible participants were identified upon presentation to the NCTLD outpatient and inpatient therapeutic departments for treatment initiation.

The R2D2 study, under which data for this analysis was collected, was approved by research ethics committees at the University of California San Francisco (20-32670), the Medical Faculty of the University of Heidelberg (S-519/2023), and the local ethics committee of NCTLD (FWA00020831). All participants provided written informed consent. The study’s reporting conforms to Standards for Reporting of Diagnostic Accuracy Studies (STARD) guidelines (checklist attached in supplementary material).

### Xpert XDR testing

Xpert XDR implementation in Georgia as a decentralized follow-on test to Xpert MTB/RIF Ultra for patients with confirmed rifampicin-resistant TB started from July 2022 at NCTLD, followed by country-wide implementation from September 2022. Testing was done by trained laboratory technicians or healthcare workers in primary care centres and zonal laboratories. Xpert XDR results were recorded as MTB detected, MTB not detected or invalid, and resistance detection per drug as resistance detected, not detected, indeterminate or invalid. If no MTB was detected in a sample, Xpert XDR showed invalid for resistance detection results. Indeterminate results indicated that resistance could not definitively be assessed by the assay algorithm, whereas an invalid result was indicating that the sample processing control failed. We did not record repeat test results.

### Microbiological testing

All eligible, consented patients provided sputum at enrollment which underwent culture inoculation using mycobacterium growth indicator tube on the Bactec 960 system (MGIT, Becton Dickinson, USA) and phenotypic DST including rifampicin, isoniazid, moxifloxacin, and levofloxacin using WHO recommended critical concentrations [11]. All clinical information, including medical history and treatment profile, was captured on electronic case report forms, while results from TB testing, including Xpert XDR, mycobacterial culture, and DST, were extracted from the NCTLD laboratory information system. DNA extraction from positive culture isolates and whole-genome-sequencing (WGS) was conducted under the R2D2 protocol for a subset of participants included in this study. Details on microbiological testing can be found in the supplementary material.

### Clinical Management of DR-TB

Rifampicin-resistant TB patients were managed according to the Georgia National TB treatment guidelines following the last update of WHO treatment guideline by clinicians through routine programmatic care [12]. The R2D2 study team was not involved in decisions on treatment initiation and drug regimens. The first choice regimen was 6-month BPaL(M) (bedaquiline, pretomanid, linezolid, optional moxifloxacin). An alternative 6-9 months all-oral modified short treatment regimen included combinations of bedaquiline, moxifloxacin, clofazimine, pyrazinamide, ethambutol, isoniazid, ethionamide, and linezolid. Additionally, an 18-month individualized regimen was available for use based on pDST results (e.g., for those with resistance to bedaquiline or other key second-line drugs).

### Outcomes

The primary outcome was diagnostic accuracy of routine programmatic Xpert XDR testing for detection of isoniazid and fluoroquinolone resistance compared to a phenotypic DST reference standard. Secondary outcomes included: (i) diagnostic accuracy of Xpert XDR compared to pDST according to intention-to-diagnose; (ii) diagnostic accuracy of Xpert XDR compared to a composite genotypic and phenotypic reference standard; (iii) diagnostic accuracy for low-level fluoroquinolone resistance detection (based on pDST at the WHO-recommended clinical breakpoint) [11]; (iv) time to DR-TB treatment initiation after Xpert XDR testing; (v) proportion of patients started on appropriate DR-TB treatment regimen(s) based on Xpert XDR results.

### Statistical Analysis

We calculated sensitivity, specificity, positive predictive value (PPV), and negative predictive value (NPV) in respect to the reference standard with binomial 95% confidence intervals. Observations with indeterminate and invalid results of both the index and reference standard tests were excluded from primary diagnostic accuracy analyses. Indeterminate rates for the index test were calculated, and we performed an intention-to-diagnose analysis in which we considered indeterminate/invalid index test results as negative for resistance [13, 14]. We performed diagnostic accuracy analyses using a composite reference standard of pDST and WGS for a subset of participants with available WGS data (see Supplementary Methods). We modelled predictive values for fluoroquinolone resistance detection across a range of resistance prevalences (5%-30%) relevant to high DR-TB burden settings. Time to treatment was calculated as time interval in days between Xpert XDR testing and treatment initiation, and reported using the median and interquartile range (IQR). Turn-around time for Xpert XDR was calculated using the date of Xpert Ultra testing as a proxy for sputum collection. The proportion of participants initiated on appropriate treatment was based on appropriate use of fluoroquinolone, i.e. inclusion of fluoroquinolone in treatment regimen if Xpert XDR demonstrated fluoroquinolone sensitivity, and exclusion of fluoroquinolone from the regimen if Xpert XDR showed resistance. An alluvial diagram was used to visualize the relation between index and reference test results and initiated fluoroquinolone treatment. Analyses were conducted using Stata/SE 18.0 (StataCorp, College station, Texas) and R Version 4.3.2 (The R Foundation for Statistical Computing).

## Results

### Baseline characteristics

201 patients were screened for eligibility for the R2D2 TB Network DR-TB study. 28 (13.9%) patients were excluded due to negative or contaminated sputum culture results and 3 patients were missing culture at the time of analysis (**Error! Reference source not found**.). 30 participants were excluded from accuracy analysis due to missing Xpert XDR testing (30/173, 17.3%) but included in clinical utility analyses, and 140 were included in the accuracy analysis. 38/140 were female, the median age was 45 years, and 30 (21.4%) and 15 (10.7%) stated they had previously been treated for drug susceptible TB (DS-TB) and DR-TB, respectively. 129/140 (94.9%) participants had isoniazid resistance, and 46/140 (33.8%) participants were resistant to fluoroquinolone by the primary reference standard (Table 1).

**Table 1:** Baseline Characteristics.

| N | Total<br>N=140 | fluoroquinolone-<br>resistant<br>N=46 | No Xpert MTB/XDR<br>N=30 |
| --- | --- | --- | --- |
| **Sex** |  |  |  |
| Male | 102 (72.9%) | 36 (78.3%) | 27 (90.0%) |
| Female | 38 (27.1%) | 10 (21.7%) | 3 (10.0%) |
| **Age, years (SD)** | 46 (14.9) | 42 (13.5) | 49 (15.1) |
| **BMI, median (IQR)** | 20.4 (18.4-23.1) | 20.6 (19.1-22.6) | 21.1 (18.1-23.4) |
| **Comorbidities (self-report)** |  |  |  |
| Living with HIV (PLHIV)<sup>a</sup> | 5 (3.6%) | 2 (4.3%) | 2 (6.7%) |
| CD4 count in PLHIV, mean (SD) | 271 (223.6) | 65 (18.4) | 74 (0.0) |
| Living with diabetes<sup>b</sup> | 8 (5.7%) | 1 (2.2%) | 3 (10.0%) |
| **Symptoms (in the past 30 days)** |  |  |  |
| Cough | 132 (94.3%) | 44 (95.7%) | 28 (93.3%) |
| Night Sweats | 123 (87.9%) | 42 (91.3%) | 21 (70.0%) |
| Weight Loss | 101 (72.1%) | 32 (69.6%) | 17 (56.7%) |
| Fever | 96 (68.6%) | 32 (69.6%) | 17 (56.7%) |
| **DR-TB contact** |  |  |  |
| No | 73 (52.1%) | 25 (54.3%) | 13 (43.3%) |
| Yes | 30 (21.4%) | 10 (21.7%) | 7 (23.3%) |
| Unsure | 37 (26.4%) | 11 (23.9%) | 10 (33.3%) |
| **Previous TB treatment** | **41 (29.3%)** | **12 (29.3%)** | **9 (30.0%)** |
| Previous DR-TB treatment<sup>c</sup> | 15 (36.6%) | 3 (25.0%) | 2 (22.3%) |
| Previous DS-TB treatment<sup>d</sup> | 30 (73.2%) | 10 (83.3%) | 6 (66.7%) |
| **Semiquantitative Grade of Xpert Ultra** |  |  |  |
| Very Low/Low | 69 (49.3%) | 24 (52.2%) | 9 (30.0%) |
| Medium/High | 71 (50.7%) | 22 (47.8%) | 9 (30.0%) |
| **TB regimen started** | **135 (96.4%)** | **43 (93.5%)** | **29 (96.7%)** |
| BPaLM | 57 (42.2%) | 8 (18.6%) | 9 (31.0%) |
| BPaL | 29 (21.5%) | 28 (65.1%) | 4 (13.8%) |
| Short all-oral regimen<sup>e</sup> | 42 (31.1%) | 5 (11.6%) | 13 (44.8%) |
| Other regimen | 7 (5.2%) | 2 (4.7%) | 3 (10.3%) |
| **Isoniazid resistance prevalence<sup>f</sup>** | 129 (94.9%) | 45 (97.8%) | 23 (92.0%) |
| **Fluoroquinolone resistance prevalence<sup>f</sup>** | 46 (33.8%) | - | 8 (32.0%) |

<sup>a</sup>3 missing <sup>b</sup>1 missing <sup>c</sup>2 missing <sup>d</sup>3 missing <sup>e</sup>Bedaquiline, Fluoroquinolone, Linezolid, Clofazimine, Cycloserine <sup>f</sup>3 exclusions (unavailable pDST). DS-TB drug sensitive TB ; DR-TB drug resistant TB ; SD standard deviation ; IQR inter-quartile range; BPaLM Bedaquiline, Pretomanid, Linezolid and Moxifloxacin ; BPaL Bedaquiline, Pretomanid, Linezolid

### Xpert XDR

Xpert XDR sensitivity for isoniazid resistance was 99.2% (95% CI, 95.5%-100.0%, n=128) and specificity was 100% (95% CI, 54.1%-100%) compared to phenotypic DST. Sensitivity and specificity for fluoroquinolone resistance were 88.4% (95%CI, 74.9%-96.1%, n=125) and 100.0% (95%CI, 95.6%-100%, Table 2), respectively. Diagnostic accuracy estimates did not differ by sex or Xpert Ultra semiquantitative grades, although fluoroquinolone sensitivity was lower in those with high or medium semiquantitative grade (77.3% for medium/high compared to 100% for low/very low, p=0.02, Supplementary table 1). Accuracy was also similar in secondary analysis using the composite refence standard (Supplementary table 2).

**Table 2:**
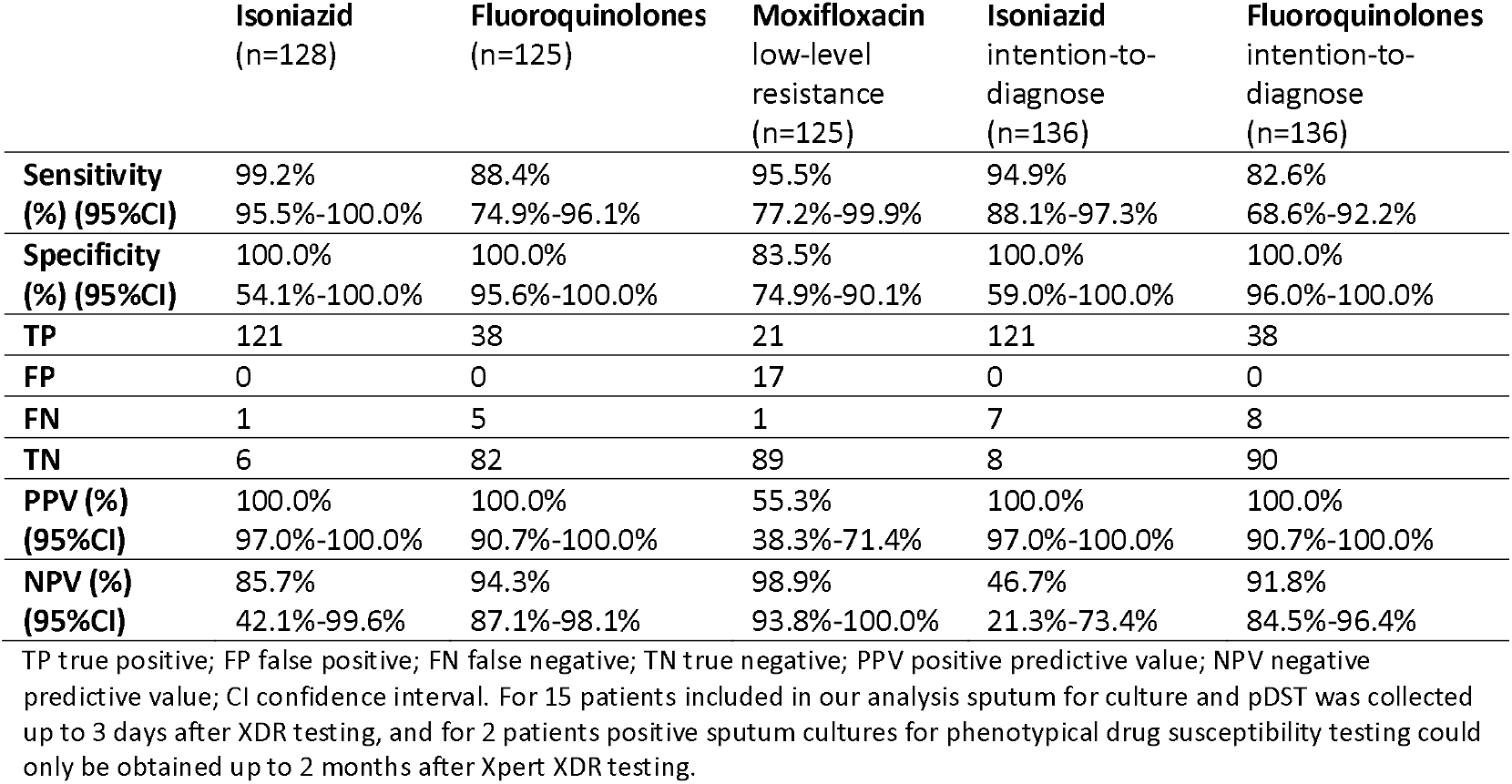
Diagnostic accuracy of Xpert XDR compared to phenotypic drug susceptibility testing.

No invalid results were observed for MTB detection. Invalid results for resistance detection only occurred in samples where no MTB was detected by Xpert XDR. No indeterminate results were registered for isoniazid resistance detection. 3.8% (5/131) of results for fluoroquinolone resistance were indeterminate, occurring both in samples with high and low bacillary load (Supplementary table 3). Intention-to-diagnose analysis found a lower sensitivity for isoniazid resistance (94.9%) and fluoroquinolone resistance detection (82.6%) compared to the primary analysis (Table 2, Supplementary tables 4 and 5). Including indeterminate-invalid results, 17.4% (8/46) of patients with phenotypic fluoroquinolone resistance and 6.4% (8/129) of patients with phenotypic isoniazid resistance were missed by Xpert XDR (Supplementary 4 and 5). Xpert XDR showed consistently high PPV and NPV for isoniazid and fluoroquinolone resistance detection across a range of resistance prevalence rates (Supplementary table 6).

**Table 3:** Clinical use-cases of Xpert XDR across settings.

| Use case | Guide choice of WHO BDQ-containing 6 month regimen (BPALM and BPAL) | Guide drug choice of 9 months all-oral regimen | Guide drug choice of longer, individualised DR-TB regimen | Detection of INH mono-resistance | Guide treatment decisions for patients without viable culture / replace programmatic pDST for INH and FQ |
| --- | --- | --- | --- | --- | --- |
| **Xpert XDR result** | 1. FQ resistance | 1. INH resistance<br>2. Low-level INH resistance<br>3. FQ resistance<br>4. Low-level FQ resistance | 1. INH resistance<br>2. FQ resistance<br>3. Low-level FQ resistance<br>4. ETO resistance<br>5. AMK resistance | 1. INH resistance<br>2. FQ resistance<br>3. Low-level INH resistance | 1. INH resistance<br>2. FQ resistance |
| **Outcome based on Xpert XDR result** | 1. Exclude MFX/FQ (i.e. BPAL instead of BPALM) | 1. Exclude INH<br>2. Consider high-dose INH<br>3. Exclude fluoroquinolones<br>4. Consider high-dose moxifloxacin (and confirmatory pDST at 1.0mg/L) | 1. Exclude INH<br>2. Exclude FQ<br>3. Consider high-dose MFX (and confirmatory pDST at 1.0mg/L)<br>4. Exclude ETO<br>5. Exclude AMK | 1. Need for addition of FQ to regimen<br>2. Exclusion of FQ in regimen<br>3. Consideration of high-dose isoniazid | 1. Selection of DS-TB/DR-TB regimens |
| **Advantages** | + avoid toxicity and costs of ineffective drug treatment<br>+ avoid increased risk of acquired drug resistance [20] | + avoid toxicity and costs of ineffective drug treatment<br>+ could reduce need for pDST for (e.g. INH, FQ)<br>+ rapid treatment initiation<br>+ avoid increased risk of acquired drug resistance [20] | + avoid toxicity and costs of ineffective drug treatment<br>+ could reduce need for pDST for (e.g. INH, FQ)<br>+ rapid treatment initiation with confidence around susceptibility/resistance to INH, FQ, ETO and injectables | + avoid ineffective drug treatment | + enable resistance testing and informed treatment decisions for patients without culture<br>+ cost saving |
| **Disadvantages** | ~10% started on FQ inappropriately due to missed FQ resistance |  |  | Expensive to use for all TB, will depend on background INH mono-resistance rate | Missing some INH and FQ resistance<br>Reduced surveillance for new |
|  | Lack of information on BDQ and LZD susceptibility |  |  |  | mutations |

BDQ Bedaquiline , INH isoniazid , FQ fluoroquinolones , ETO ethionamide, AMK amikacin, LZD linezolid

Xpert MTB XDR detected MTB in 80.0% (12/15) of samples that had negative or contaminated cultures, 58.4% (7/12) of these showed isoniazid resistance, and 8.3% (1/12) showed fluoroquinolone resistance. Xpert XDR failed to detect MTB in 9/140 (6.4%) culture-positive samples, all had low or very low semiquantitative grades on Xpert Ultra, 77.8% (7/9) were isoniazid resistant by phenotypic DST and 22.2% (2/9) were fluoroquinolone resistant by phenotypic DST.

### Clinical utility

Median turn-around time for Xpert XDR was 1 day (IQR 1-3) and median time from Xpert XDR result to treatment was 4 days (IQR 1-7). The median time between sputum collection and pDST results was 40 days (IQR 27-59). Data on fluoroquinolone treatment was available for 135/140 participants with Xpert XDR results. 115/121 (95.0%, 95%CI 89.5-98.2%) participants had fluoroquinolones appropriately prescribed based on their Xpert XDR result. 98.8% (85/86, 95%CI 93.7-100.0%) of those with fluoroquinolone-sensitive Xpert XDR results had them prescribed, while 85.7% (30/35, 95%CI 69.7-95.2%) of those with fluoroquinolone-resistance on Xpert XDR had no fluoroquinolone prescribed. 91.7% (110/120 95%CI 85.2-95.9%) of participants with valid Xpert XDR and pDST results had fluoroquinolones appropriately prescribed based on reference standard results, compared to 82.9% (29/35. 95%CI 66.4-93.4%, p=0.13) of participants without valid Xpert XDR results 4.1% (5/121) of participants initiated on DR-TB treatment were started on fluoroquinolone treatment despite Xpert XDR indicating resistance, all of whom showed phenotypic resistance to fluoroquinolone. For 3/5 patients, fluoroquinolones were removed from the regimen before pDST results became available. Figure 3 shows the relationship between Xpert XDR results, initiation of a fluoroquinolone treatment, and phenotypic resistance for fluoroquinolone in patients with and without valid Xpert XDR results.

**Figure 1:**
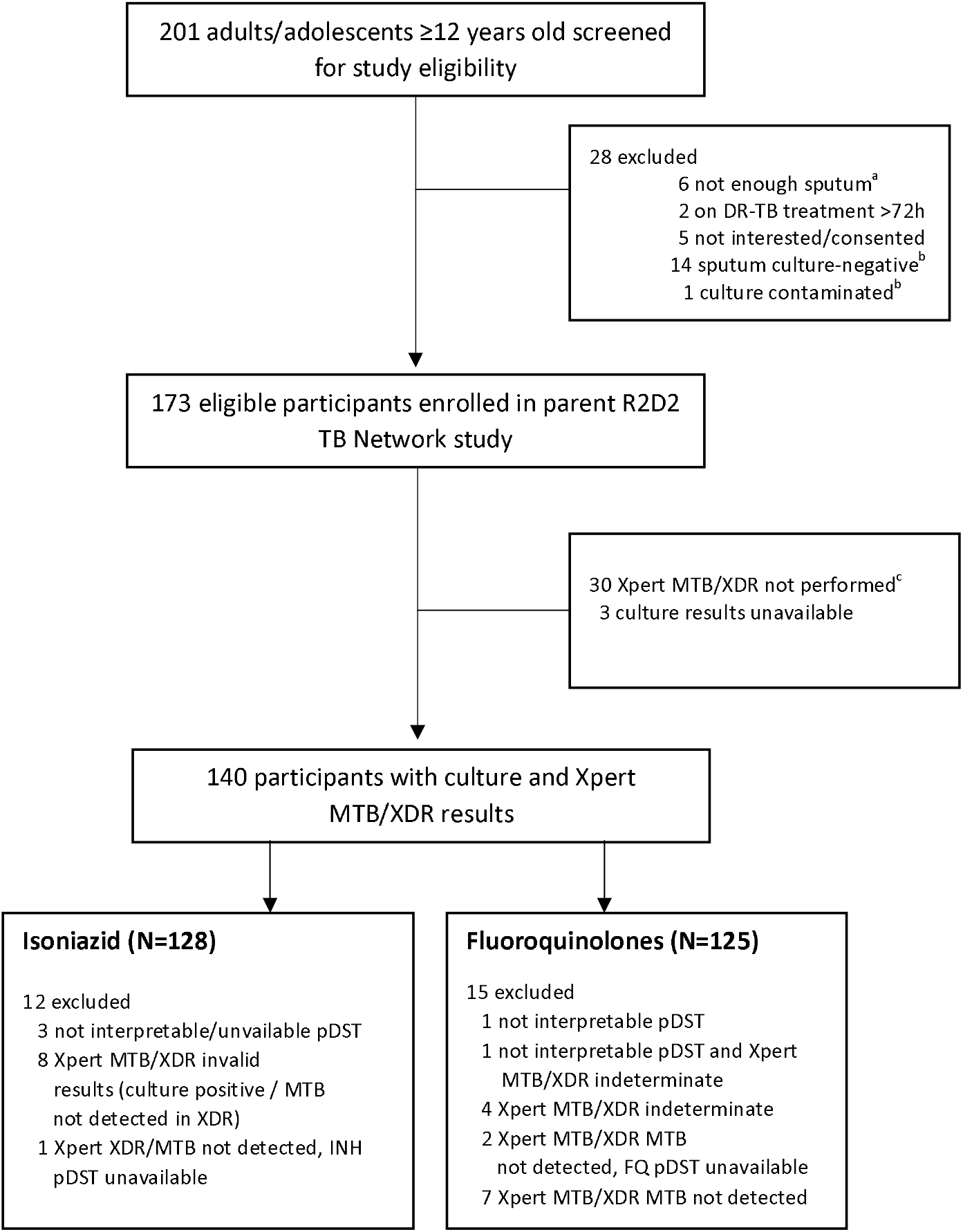
Participant Flow. ^a^A minimum of 6ml of sputum was required for enrollment in the R2D2 TB Network study ^b^late exclusions ^c^excluded from accuracy analyse but included in clinical utility analyses. DR-TB Drug resistant TB, INH Isoniazd, FQ Fluoroquinolone, pDST phenotypic drug susceptibility testing

**Figure 3:**
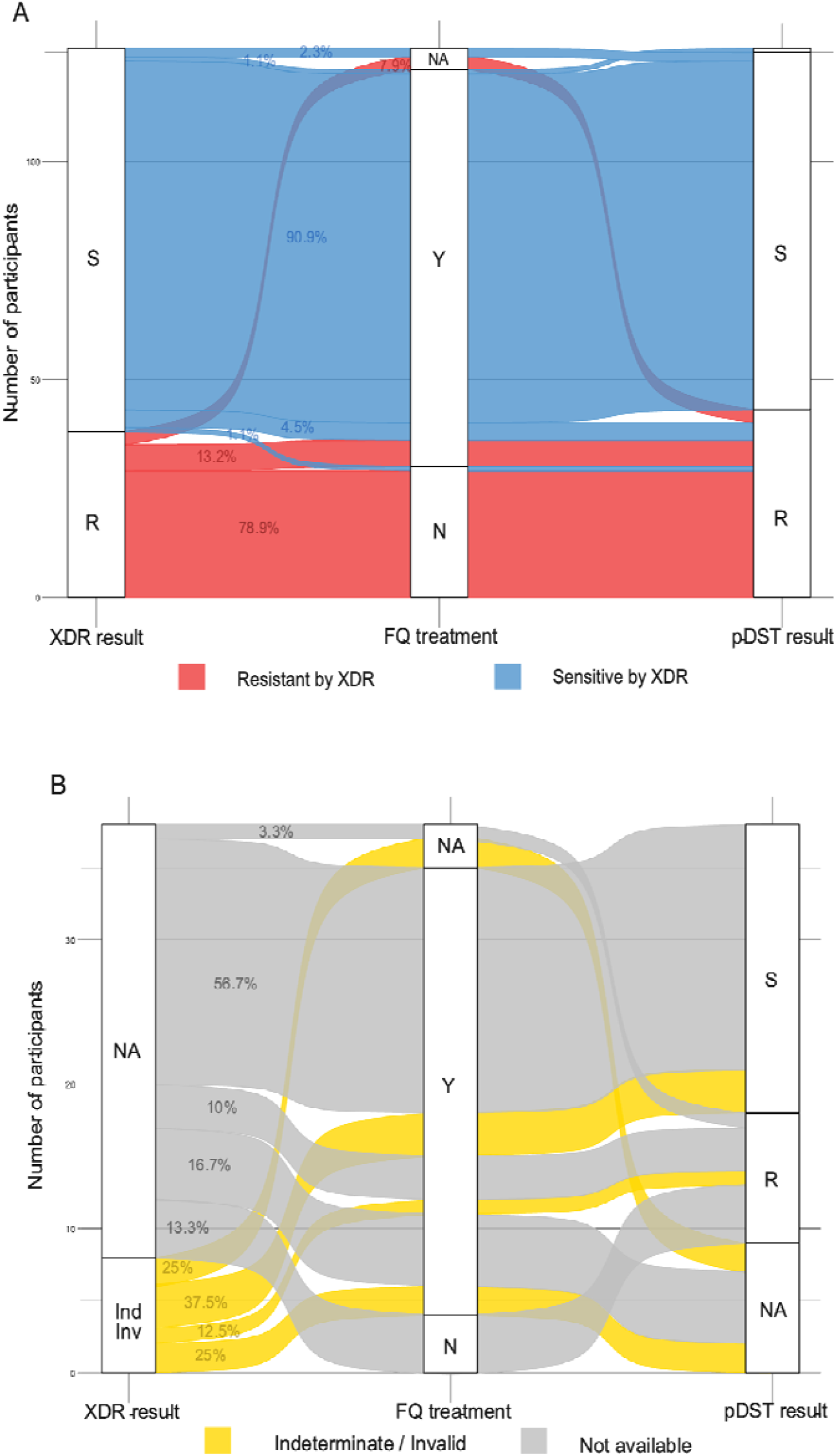
Relation between Xpert XDR results, fluoroquinolone treatment, and phenotypic DST results in patients with (A) and without (B) valid Xpert MTB/XDR results. NA not available (missing, not done), Ind indeterminate, Inv Iivalid, Y yes, N no, S sensitive, R resistant, FQ treatment fluoroquinolone treatment

## Discussion

In this pragmatic diagnostic accuracy evaluation, we found that Xpert XDR has high sensitivity and specificity for both isoniazid and fluoroquinolone resistance when implemented in a programmatic setting. The PPV and NPV for both drugs were consistently high across a range of prevalences. Indeterminate resistance results from Xpert XDR were uncommon (4% for fluoroquinolones), and mostly associated with low sample mycobacterial load. The median time to treatment after Xpert XDR testing was four days. Phenotypic DST took an average of six weeks longer than Xpert XDR. The use of fluoroquinolone was largely consistent with Xpert XDR results. While a small subset of sensitive results (1%) for fluoroquinolone were later determined resistant on phenotypic DST, there were no false positive resistance results on Xpert XDR. Nevertheless, some participants were initiated on a fluoroquinolone containing regimen despite Xpert XDR indicated fluoroquinolone resistance.

Xpert XDR demonstrated similar diagnostic accuracy for isoniazid and fluoroquinolone resistance in routine care compared to early accuracy clinical studies [6, 7, 15, 16]. However, our results of sensitivity for fluoroquinolone resistance detection (88%, 83% in intention-to-diagnose analysis) fall just below the recent WHO TPP targets (>90%) [17], and are slightly lower than estimates from testing programmatic specimens in South Africa [18]. Surprisingly, we observed lower sensitivity for fluoroquinolone resistance in samples with higher bacillary burden based on Xpert Ultra semi-quantative grade. Previous accuracy studies have reported no difference in sensitivity by smear status [6], and further data on this will be useful given our relatively small sample size. As previously demonstrated in research settings, Xpert XDR also exhibits a low indeterminate rate for isoniazid resistance detection with programmatic use [15, 19]. In this real-world setting, Xpert XDR enabled early informed treatment decisions for patients with rifampicin-resistant TB, with treatment initiation within a median of 4 days of testing, substantially surpassing the turn-around time of phenotypic DST by weeks. Although missing approximately 10% of fluoroquinolone resistance, the high specificity of Xpert XDR for fluoroquinolone resistance, together with its fast turn-around time, and the limitations of culture-based fluoroquinolone resistance testing, Xpert XDR has significant utility to assess eligibility for inclusion of moxifloxacin in the WHO recommended BPaLM regimen [12].

Prior to implementation and scale-up of a new rapid DST, it is essential to clearly define the appropriate clinical use cases, and how results should be interpreted by clinicians to inform patient treatment. Use cases proposed for Xpert XDR as a rapid molecular DST for isoniazid and fluoroquinolones include as a reflex test following detection of rifampicin resistance [7, 16]. Based on current WHO-treatment guidelines, previous literature, and our results of programmatic accuracy and utility, we propose several potential use cases for Xpert XDR (Table 3) [7, 12, 16, 18].

Of note for patients with rifampicin-resistant TB, the appropriate use of moxifloxacin in the BPaL(M) regimen is not only important for avoiding unnecessary side effects caused by a non-effective drug, but also when considering the risk of acquiring further drug resistance on some regimens in the context of undiagnosed fluoroquinolone resistance [20]. There are also concerns about bedaquiline resistance and the use of the BPaL regimen. However, the detection of fluoroquinolone resistance through Xpert XDR could prompt expedited testing for bedaquiline resistance through targeted next generation sequencing and/or phenotypic DST, if available [16]. Additionally, in our study, 80% of culture negative samples had positive XDR results, yielding important information on drug resistance that would otherwise not be available.

5% of patients included in this analysis were started on a treatment regimen that included ineffective fluoroquinolone, and clinical data showed that treatment was changed for most when phenotypic results became available. This could be due to several factors, including unfamiliarity with Xpert XDR or the diagnostic algorithm incorporating its use or suboptimal communication of results.

While the assay has demonstrated high diagnostic accuracy for isoniazid and fluoroquinolone resistance detection in research as well as routine care settings, there are limitations to its use in DR-TB due to implementation of newer, bedaquiline-based regimens. Xpert XDR’s resistance panel, while useful for determining fluoroquinolone resistance prior to starting BPaL(M), and the use of ethionamide and amikacin in the longer regimens, does not align well with the current DR-TB treatment recommendations, most importantly by lacking resistance targets for bedaquiline [12]. According to the recent WHO TPP, rapid molecular DSTs should at least include targets for rifampicin, isoniazid, fluoroquinolone, as well as bedaquiline, to help protect acquisition of resistance to newer second-line DR-TB drugs [17].

Another limitation to the widespread use and the assay’s potential clinical impact is the cost of the cartridge, currently around US$15 for Global Fund, Stop TB Partnership, USAID, and Cepheid Global Access Program. Considering that an additional test is needed to determine resistance to rifampicin, the cost of Xpert XDR does not meet minimal TPP price requirements for DST [17]. Also the 10-colour GeneXpert platform required to run Xpert XDR may not be available across settings. The potential of the test when used in a real-world programmatic setting demonstrated in our analysis supports the calls for an affordable pricing of the cartridge to take down the costs of comprehensive drug susceptibility testing. However, Xpert XDR may be a cost-effective replacement or alternative for phenotypic DST in some settings, given the high running costs of biosafety-level 3 laboratories needed for mycobacterial culture.

The main strengths of this study are its embeddedness in a real-world programmatic setting with robust data collection and quality control for microbiological testing within a large diagnostic accuracy trial and the availability of clinical patient-level data to relate testing results to treatment decision making. We have also performed an intention-to-diagnose analysis to accurately demonstrate diagnostic performance in a real-world setting. However, the programmatic nature of our data comes with some limitations, including some missing data on date of sputum collection, and a small number of patients with delays between sputum samples used for Xpert XDR and those used for phenotypic DST. Lastly, isoniazid resistance prevalence is extremely high in the included patient population, thereby our sample size for specificity of isoniazid resistance detection analyses is small, resulting in wide confidence intervals for specificity estimates.

## Conclusion

The high accuracy of Xpert XDR for detection of isoniazid and fluoroquinolone resistance in a programmatic setting is in line with estimates from diagnostic accuracy studies, although below WHO TPP targets, expecially when accounting for indeterminate/invalid results. In a setting with available infrastructure and resources for pDST, the added benefit of Xpert XDR testing for clinical decision making lies mainly in a substantially shorter turn-around time. However, a key-use case for the assay might be in settings without access to reliable pDST. While Xpert XDR remains useful as one of few endorsed rapid, low complexity molecular tests for isoniazid and fluoroquinolone resistance, there is a strong need for rapid DST that includes bedaquiline and other newer TB drugs, as well as the need for assessment of impact against diagnostic guidelines and algorithms.

## Supporting information

supplementary methods and results

## Data Availability

All data produced in the present study are available upon reasonable request to the authors

## Acknowledgments

AG-W, NT, and TP conceptualized and designed the study. Funding for the R2D2 project under which data collection for this study was conducted was acquired by CMD and NT. NT, NM, AT, MG, NB, and TP conducted data collection, AG-W, NT, TP coordinated the research activity. CMD, AG-W, and NT provided scientific support. TP and AG-W did the formal data analysis. TP, AG-W, and NT wrote the manuscript draft. All authors had full access to all data in the study, contributed to the interpretation of results, reviewed and edited the manuscript, approved the final version and agreed to the submission for publication.

## References

1. Global tuberculosis report 2023. Geneva: World Health Organization, 2023.

2. Murray CJL, Ikuta KS, Sharara F, et al. Global burden of bacterial antimicrobial resistance in 2019: a systematic analysis. The Lancet 2022; 399(10325): 629–55.

3. Boehme CC, Nicol MP, Nabeta P, et al. Feasibility, diagnostic accuracy, and effectiveness of decentralised use of the Xpert MTB/RIF test for diagnosis of tuberculosis and multidrug resistance: a multicentre implementation study. The Lancet 2011; 377(9776): 1495–505.

4. MacLean EL-H, Miotto P, González Angulo L, et al. Updating the WHO target product profile for next-generation Mycobacterium tuberculosis drug susceptibility testing at peripheral centres. PLOS Global Public Health 2023; 3(3): e0001754.

5. WHO consolidated guidelines on tuberculois. Module 3: diagnosis - rapid diagnostics for tuberclosis detection 2021 update. Geneva: Word Health Organization, 2021.

6. Penn-Nicholson A, Georghiou SB, Ciobanu N, et al. Detection of isoniazid, fluoroquinolone, ethionamide, amikacin, kanamycin, and capreomycin resistance by the Xpert MTB/XDR assay: a cross-sectional multicentre diagnostic accuracy study. The Lancet Infectious Diseases 2022; 22(2): 242–9.

7. Pillay S, Steingart KR, Davies GR, et al. Xpert MTB/XDR for detection of pulmonary tuberculosis and resistance to isoniazid, fluoroquinolones, ethionamide, and amikacin. Cochrane Database of Systematic Reviews 2022; (5).

8. Omar SV, Louw G, Ismail F, et al. Performance evaluation of the Xpert MTB/XDR test for the detection of drug resistance to <em>Mycobacterium tuberculosis</em> among people diagnosed with tuberculosis in South Africa. medRxiv 2024: 2024.02.16.24302824.

9. Chen X, Li R, Ge S, et al. Rapid Detection of Extensive Drug Resistance by Xpert MTB/XDR Optimizes Therapeutic Decision-Making in Rifampin-Resistant Tuberculosis Patients. Journal of Clinical Microbiology 2023; 61(6): e01832–22.

10. Rapid Research in Diagnostics Development for TB Network (R2D2 TB Network) Study. In: University Hospital H, Christian Medical College VI, Vietnam National Lung H, et al., 2021.

11. Technical Report on critical concentrations for drug susceptibility testing od medicines used in the treatment of drug-resistant tuberculosis. Geneva: World Health Organization, 2018. Report No.: WHO/CDS/TB/2018.5.

12. WHO consolidated guidelines on tuberculosis. Modul 4: treatment-rug-resistant tuberculosis treatment 2022 update. Geneva: World Health Organization, 2022.

13. Stahlmann K, Reitsma JB, Zapf A. Missing values and inconclusive results in diagnostic studies – A scoping review of methods. Statistical Methods in Medical Research 2023; 32(9): 1842– 55.

14. Schuetz GM, Schlattmann P, Dewey M. Use of 3×2 tables with an intention to diagnose approach to assess clinical performance of diagnostic tests: meta-analytical evaluation of coronary CT angiography studies. BMJ : British Medical Journal 2012; 345: e6717.

15. Maya T, Wilfred A, Lubinza C, et al. Diagnostic accuracy of the Xpert® MTB/XDR assay for detection of Isoniazid and second-line antituberculosis drugs resistance at central TB reference laboratory in Tanzania. BMC Infectious Diseases 2024; 24(1): 672.

16. Saluzzo F, Masood F, Batignani V, et al. TB drug susceptibility testing in high fluoroquinolone resistance settings. IJTLD Open 2024; 1(5): 230–5.

17. Target product profiles for tuberculosis diagnosis and detection of drug resistance. Geneva: World Health Organization, 2024.

18. Centner CM, Munir R, Tagliani E, et al. Reflex Xpert MTB/XDR Testing of Residual Rifampicin-Resistant Specimens: A Clinical Laboratory-Based Diagnostic Accuracy and Feasibility Study in South Africa. Open Forum Infectious Diseases 2024; 11(8).

19. Sethi S, Sharma S, Aggarwal AN, Dhatwalia SK, Rana R, Yadav R. Xpert MTB/XDR assay: rapid TB drug resistance detection. Infection 2024.

20. Mallick JS, Nair P, Abbew ET, Van Deun A, Decroo T. Acquired bedaquiline resistance during the treatment of drug-resistant tuberculosis: a systematic review. JAC-Antimicrobial Resistance 2022; 4(2).

