## supplementary methods and results for "Programmatic diagnostic accuracy and clinical utility of Xpert MTB/XDR in patients with rifampicin-resistant tuberculosis in Georgia"

*contributed equally

Corresponding Author:

**Supplementary Methods**

Xpert XDR testing

Georgian routine diagnostic guidelines recommended Xpert XDR testing for all patients with confirmed rifampicin resistant TB by Xpert Ultra and a negative smear microscopy result from the start of programmatic implementation in July 2022 until March 2024. From April 2024 Xpert XDR testing was recommended for all patients positive for TB on Xpert Ultra regardless of rifampicin resistance and smear status. At all times clinicians had the option to request Xpert XDR testing at their own descretion regardless of guideline documentation.

Reference Standard Testing

Primary reference standard testing consisted of phenotypic DST in MGIT tubes at the WHO recommended critical concentrations for isoniazid (0.1 mg/L), levofloxacin (1.0 mg/L), and moxifloxacin (0.25 mg/L). Additionally, high-dose moxifloxacin susceptibility testing was done at the clinical breakpoint (1.0mg/L). Drug resistance was determined using the 1% proportion method [20]. Phenotypic DST was chosen as reference standard since it represents the current gold standard for drug resistance detection [21]. Participants were classified as resistant to fluoroquinolone by reference standard if isolates were resistant to levofloxacin and/or moxifloxacin. For a subset of participants with available WGS data we performed diagnostic accuracy analyses against a composite reference standard of phenotypic DST and WGS. Negative drug resistance under the composite reference standard was defined as an absence of WGS gene mutation(s) known to confer drug resistance as per WHO mutations catalogue 2023 and a pDST finding the isolate is sensitive to the drug at the concentrations mentioned above [22]. Reference standard test results and collected clinical information were not available to laboratory staff at the time of conducting Xpert XDR testing and vice versa.

**STARD Checklist**

| Section & Topic | No | Item | Reported on page # |
| --- | --- | --- | --- |
| TITLE OR ABSTRACT | | | |
|  | 1 | Identification as a study of diagnostic accuracy using at least one measure of accuracy  (such as sensitivity, specificity, predictive values, or AUC) | 1,2 |
| ABSTRACT | | | |
|  | 2 | Structured summary of study design, methods, results, and conclusions  (for specific guidance, see STARD for Abstracts) | 2 |
| INTRODUCTION | | | |
|  | 3 | Scientific and clinical background, including the intended use and clinical role of the index test | 3 |
|  | 4 | Study objectives and hypotheses | 3 |
| METHODS | | | |
| *Study design* | 5 | Whether data collection was planned before the index test and reference standard  were performed (prospective study) or after (retrospective study) | 3 |
| *Participants* | 6 | Eligibility criteria | 3,4 |
|  | 7 | On what basis potentially eligible participants were identified  (such as symptoms, results from previous tests, inclusion in registry) | 3,4 |
|  | 8 | Where and when potentially eligible participants were identified (setting, location and dates) | 4 |
|  | 9 | Whether participants formed a consecutive, random or convenience series | 3 |
| *Test methods* | 10a | Index test, in sufficient detail to allow replication | 4, supplementary material |
|  | 10b | Reference standard, in sufficient detail to allow replication | 4 + supplementary methods |
|  | 11 | Rationale for choosing the reference standard (if alternatives exist) | Supplementary methods |
|  | 12a | Definition of and rationale for test positivity cut-offs or result categories  of the index test, distinguishing pre-specified from exploratory | NA |
|  | 12b | Definition of and rationale for test positivity cut-offs or result categories  of the reference standard, distinguishing pre-specified from exploratory | NA |
|  | 13a | Whether clinical information and reference standard results were available  to the performers/readers of the index test | Supplementary methods |
|  | 13b | Whether clinical information and index test results were available  to the assessors of the reference standard | Supplementary methods |
| *Analysis* | 14 | Methods for estimating or comparing measures of diagnostic accuracy | 5 |
|  | 15 | How indeterminate index test or reference standard results were handled | 5 |
|  | 16 | How missing data on the index test and reference standard were handled | 5 |
|  | 17 | Any analyses of variability in diagnostic accuracy, distinguishing pre-specified from exploratory | 5 |
|  | 18 | Intended sample size and how it was determined | NA |
| RESULTS | | | |
| *Participants* | 19 | Flow of participants, using a diagram | 6 |
|  | 20 | Baseline demographic and clinical characteristics of participants | 7,8 |
|  | 21a | Distribution of severity of disease in those with the target condition | NA |
|  | 21b | Distribution of alternative diagnoses in those without the target condition | 8 |
|  | 22 | Time interval and any clinical interventions between index test and reference standard | 9 |
| *Test results* | 23 | Cross tabulation of the index test results (or their distribution)  by the results of the reference standard | Supplementary results |
|  | 24 | Estimates of diagnostic accuracy and their precision (such as 95% confidence intervals) | 9 |
|  | 25 | Any adverse events from performing the index test or the reference standard | NA |
| DISCUSSION | | | |
|  | 26 | Study limitations, including sources of potential bias, statistical uncertainty, and generalisability | 11 |
|  | 27 | Implications for practice, including the intended use and clinical role of the index test | 12 |
| OTHER INFORMATION | | | |
|  | 28 | Registration number and name of registry | NA |
|  | 29 | Where the full study protocol can be accessed | 3 |
|  | 30 | Sources of funding and other support; role of funders | 15 |

**Supplementary Result****s**

Supplementary table 1 Sub-group analysis for Diagnostic accuracy of Xpert MBT/XDR against pDST

|  | **Isoniazid**  (n=128) | | **Fluoroquinolones**  (n=125) | |
| --- | --- | --- | --- | --- |
| **Xpert Ultra Grade** | **Very Low/Low** | **Medium/High** | **Very Low/Low** | **Medium/High** |
| N | 59 | 69 | 56 | 69 |
| Sensitivity (%) (95%CI) | 98.3  90.6-100.0 | 100.0  94.5-100.0 | 100.0  83.9-100.0 | 77.3 54.6-92.2 |
| Specificity (%) (95%CI) | 100.0  15.8-100.0 | 100.0  39.8-100.0 | 100.0  90.0-100.0 | 100.0 92.5-100.0 |
| TP | 56 | 65 | 21 | 17 |
| FP | 0 | 0 | 0 | 0 |
| TN | 2 | 4 | 35 | 47 |
| FN | 1 | 0 | 0 | 5 |
| **Sex** | **Male** | **Female** | **Male** | **Female** |
| N | 91 | 37 | 89 | 36 |
| Sensitivity (%) (95%CI) | 100.0  95.8-100.0 | 97.2  85.5-99.9 | 88.2  72.5-96.7 | 88.9 51.8-99.7 |
| Specificity (%) (95%CI) | 100.0  47.8-100.0 | 100.0  2.5-100.0 | 100.0  93.5-100.0 | 100.0  87.2-100.0 |
| TP | 86 | 35 | 30 | 8 |
| FP | 0 | 0 | 0 | 0 |
| TN | 5 | 1 | 55 | 27 |
| FN | 0 | 1 | 4 | 1 |

TP true positive; FP false positive; TN true negative; FN false negative; CI confidence interval

Supplementary table 2 Diagnostic accuracy of Xpert XDR against a composite reference standard of pDST and WGS

|  | **Isoniazid**  (n=68) | **Fluoroquinolones**  (n=69) | **Isoniazid**  (intention-to-diagnose)  (n=69) | **Fluoroquinolones**  (intention-to-diagnose)  (n=69) |
| --- | --- | --- | --- | --- |
| Sensitivity (%) (95%CI) | 91.7%  89.8%-99.6% | 87.0%  66.4%-97.2% | 97.0%  89.6%-99.6% | 83.3%  62.6%-95.3% |
| Specificity (%) (95%CI) | 100.0%  15.8%-100.0% | 100.0%  92.1%-100.0% | 100.0%  15.8%-100.0% | 100.0%  92.1%-100.0% |
| TP | 65 | 20 | 65 | 20 |
| FP | 0 | 0 | 0 | 0 |
| FN | 1 | 3 | 2 | 4 |
| TN | 2 | 45 | 2 | 45 |
| PPV (%)  (95%CI) | 100.0%  94.5%-100.0% | 100.0%  83.2%-100.0% | 100.0%  94.5%-100.0% | 100.0%  83.2%-100.0% |
| NPV (%)  (95%CI) | 66.7%  9.4%-99.2% | 93.8%  82.8%-98.7% | 50.0%  6.8%-93.2% | 91.8%  80.4%-97.7% |
| TP true positive; FP false positive; TN true negative; FN false negative; PPV positive predictive value; NPV negative predictive value; CI confidence interval | | | | |

Supplementary table 3 Xpert XDR results by Xpert Ultra semiquantitative grade

|  | **Xpert Ultra semiquantitative Grade** | |
| --- | --- | --- |
|  | Very Low/Low | Medium/High |
| N | 69 (100.0%) | 71 (100.0%) |
| **MTB detection** | | |
| MTB not detected | 9 (13.0%) | 0 (0.0%) |
| MTB detected | 60 (87.0%) | 71 (100.0%) |
| **Isoniazid** |  |  |
| Sensitive | 3 (4.4%) | 4 (5.6%) |
| Resistant | 57 (82.6%) | 67 (94.4%) |
| Invalid | 9 (13.0%) | 0 (0.0%) |
| **Fluoroquinolone** |  |  |
| Sensitive | 36 (53.2%) | 52 (73.2%) |
| Resistant | 21 (30.4%) | 17 (23.9%) |
| Indeterminate | 3 (4.4%) | 2 (2.8%) |
| Invalid | 9 (13.0%) | 0 (0.0%) |
| MTB Mycobacteriuem tuberculosis | | |

Supplementary table 4 3x3 Table Xpert XDR and pDST for Isoniazid resistance detection

| **Xpert MTB/XDR** | **pDST** | | |
| --- | --- | --- | --- |
|  | Sensitive | Resistant | Indeterminate |
| Sensitive | 6 | 1 | 0 |
| Indeterminate/Invalid | 1 | 7 | 0 |
| Resistant | 0 | 121 | 2 |
| pDST phenotypic drug susceptibility testing | | | |

Supplementary table 5 3X3 Table Xpert XDR and pDST for Fluoroquinolone resistance detection

| **Xpert MTB/XDR** | **pDST** | | |
| --- | --- | --- | --- |
|  | Sensitive | Resistant | Indeterminate |
| Sensitive | 82 | 5 | 1 |
| Indeterminate/Invalid | 8 | 3 | 1 |
| Resistant | 0 | 38 | 0 |
| pDST phenotypic drug susceptibility testing | | | |

Supplementary table 6 PPV and NPV of Xpert XDR for isoniazid and fluoroquinolone resistance detection at varying prevalence estimates

| **Resistance Prevalence** | **PPV**(%) (95%CI) | **NPV**(%) (95%CI) |
| --- | --- | --- |
| **Isoniazid** |  |  |
| 6.7% | 100%  - | 99.9%  99.6-100.0% |
| 10% | 100.0%  - | 99.9%  99.4%-100.0% |
| 20% | 100.0%  - | 99.8%  98.6%-100.0% |
| **Fluoroquinolones** |  |  |
| 5% | 100.0%  - | 99.4%  98.6%-99.7% |
| 10% | 100.0%  - | 98.7%  97.1%-99.4% |
| **Moxifloxacin CB** |  |  |
| 5% | 23.3% 16.3%-32.2% | 99.7% 98.1%-100.0% |
| 10% | 39.1%  29.2%- 50.0% | 99.4%  96.0%- 99.9% |
| PPV positive predictive value; NPV negative predictive value;  CB clinical breakpoint | | |
